# Association of biological sex with clinical outcomes following STA-MCA bypass in atherosclerotic cerebrovascular disease

**DOI:** 10.64898/2026.07.17.26358368

**Authors:** Mohammad Asif Iqbal, Joan Alsolivany, Kiarash Ferdowssian, Robert Mertens, Erin Dirk Sprünken, Lars Wessels, Peter Vajkoczy, Güliz Acker, Nils Hecht

**Author notes:** Corresponding Author: Name: Nils Hecht, MD, Current Institution: Department of Neurosurgery, Charité – Universitätsmedizin Berlin, Hindenburgdamm 30, 12203 Berlin, Germany. These authors contributed equally (Güliz Acker, Nils Hecht).

## Abstract

**Background:** Sex differences in cerebrovascular disease are established determinants of outcome in acute stroke care and vascular interventions, but evidence in cerebrovascular bypass surgery remains limited. This study examined whether biological sex was associated with outcome after superficial temporal artery to middle cerebral artery (STA-MCA) bypass in patients with atherosclerotic cerebrovascular disease (ACVD).

**Methods:** We retrospectively screened adults undergoing extracranial-to-intracranial (EC-IC) bypass (2012–2025) and included ACVD patients treated by STA-MCA bypass with available follow-up. The primary outcome was modified Rankin Scale (mRS) at latest follow-up, analyzed using proportional odds regression. Multivariable models adjusted for age, preoperative mRS, and vascular comorbidities. Cerebrovascular reserve capacity (CVRC) was analyzed in a subgroup.

**Results:** A total of 140 patients (30.7% female) were included. Disease morphology varied by sex, with more multivessel (65.1% vs. 47.4%) and stenotic disease (39.5% vs. 20.6%) in females and more isolated internal carotid artery occlusion in males (43.3% vs. 16.3%). The 30-day risk of symptomatic ischemic stroke was higher in females than in males (9.3% vs. 1.0%). A similar pattern was observed at follow-up (median 13.5 months), with ischemic events predominating in females (16.3% vs. 7.2%) and hemorrhagic events occurring exclusively in males (5.2%). Female sex was independently associated with worse functional outcome (OR 2.59, 95% CI 1.28–5.30, p=0.008). Preoperative mRS was the strongest determinant of outcome (OR 4.30, 95% CI 3.07–6.18, p<0.001). Adjusted analysis detected no significant association between CVRC and outcome (OR 0.80, 95% CI 0.24–2.70, p=0.721).

**Conclusions:** Female sex was independently associated with worse functional outcome after STA-MCA bypass, independent of preoperative functional status, hemodynamic impairment and cardiovascular comorbidities. These findings identify sex as a clinically relevant determinant of outcome in cerebrovascular bypass surgery and should be considered in future risk stratification and trial design.

## INTRODUCTION

Sex differences in cerebrovascular disease represent an increasingly recognized determinant of clinical outcome across the spectrum of acute stroke care and vascular intervention^1–3^. In observational stroke cohorts, female sex has been associated with less favorable functional outcomes after intravenous thrombolysis and mechanical thrombectomy^4,5^. It has been examined as a potential modifier of procedural risk in carotid revascularization, where several studies have identified higher perioperative complication rates in women, particularly in relation to endovascular treatment strategies^3,6,7^. These observations highlight the growing recognition of sex as a clinically relevant variable across both medical and interventional treatment paradigms. In the broader vascular surgical literature, sex-related differences in both perioperative and long-term outcomes have been documented across multiple intervention types^8–11^.

In cerebrovascular bypass surgery, sex-specific functional outcome data remain largely unexplored. This is particularly relevant given the distinct pathophysiological context and patient selection frameworks associated with extracranial–intracranial (EC-IC) revascularization. The only dedicated sex-stratified analysis of EC-IC bypass was conducted in a mixed-etiology cohort and assessed bypass patency as its primary endpoint, without reporting functional outcomes^12^. Randomized evidence from COSS and, more recently, CMOSS has shaped contemporary indications for EC-IC bypass in patients with hemodynamic cerebral ischemia. While neither trial was designed to evaluate sex as an effect modifier, both substantially advanced patient selection for contemporary bypass surgery^13,14^. Against this background, the present study investigates sex-specific determinants of functional outcome after superficial temporal artery to middle cerebral artery (STA-MCA) bypass in patients with atherosclerotic cerebrovascular disease (ACVD).

## METHODS

This study was designed as a single-center retrospective cohort study and conducted in accordance with the Declaration of Helsinki and its later amendments. The study was approved by the ethics committee of Charité – Universitätsmedizin Berlin covering both a retrospective (EA2/139/12) and a prospective registry (EA2/178/18). The requirement for written informed consent was waived due to the retrospective nature of the analysis. All consecutive adult patients (≥18 years) who underwent EC-IC bypass at the neurosurgical department of Charité – Universitätsmedizin Berlin between February 2012 and March 2025 were screened. Patients with steno-occlusive cerebrovascular disease (1) attributable to atherosclerosis who underwent (2) direct STA-MCA bypass and had (3) available follow-up data were included. Patients treated for (1) non-atherosclerotic indications or with (2) incomplete follow-up were excluded.

### Patient characteristics

Medical records were reviewed for demographic data, onset symptoms, comorbidities, medications, perioperative course, and follow-up findings. Biological sex was obtained from the medical record. Age was defined as age at time of surgery. The onset symptoms were classified as either a single ischemic event or recurrent ischemic events, which included transient ischemic attacks, clinically manifest stroke, and imaging-defined silent infarctions. Comorbidities and cardiovascular risk factors were recorded based on documented medical history and preoperative anesthesia protocol. These included hypertension, diabetes mellitus, hyperlipidemia, smoking history, chronic obstructive pulmonary disease, atrial fibrillation, chronic kidney disease, and coronary artery disease. Medication status, including antiplatelet therapy, statin use, and anticoagulation, was also documented.

### Radiological workup and hemodynamic assessment

Preoperative imaging included magnetic resonance imaging (MRI), digital subtraction angiography (DSA), and perfusion imaging for assessment of cerebrovascular reserve capacity (CVRC). Hemodynamic assessment was performed using acetazolamide-stimulated [⁹⁹ᵐTc]Tc-HMPAO single-photon emission computed tomography (SPECT) or [¹⁵O]H₂O positron emission tomography (PET). Routine computed tomography (CT) with CT angiography was performed on postoperative day one to assess patency and exclude complications, while additional imaging was obtained in case of neurological deterioration. If CT angiography was inconclusive, DSA was performed before discharge and routinely at 12 months.

### Surgical technique and perioperative management

Patients were anesthetized with propofol and remifentanil, with intraoperative mean arterial pressure targeted at 90–100 mmHg and arterial PaCO₂ maintained between 38 and 42 mmHg. The standard pedicled STA-MCA bypass was performed as previously described^15,16^. The STA branch with greater diameter was selected as the donor, while a cortical M4 branch of the MCA at the distal end of the sylvian fissure corresponding to the target point as described by Peña et al. was selected as the recipient based on diameter and flow as previously described^16,17^. The anastomosis was performed end-to-side in continuous or interrupted fashion using 10-0 monofilament, non-absorbable synthetic suture with fishmouthing of the donor vessel. Inspired oxygen fraction was maintained at 1.0 (100%) during anastomosis. Postoperatively, systolic blood pressure was targeted 10% above the individual patient’s regular systolic blood pressure for the first 12–24 hours in the neurointensive care unit.

### Follow-up and outcome assessment

The 30-day stroke rate was defined as the proportion of patients experiencing a new symptomatic ischemic stroke within 30 days of surgery, confirmed by imaging. Beyond the perioperative period, patients were clinically evaluated at 3 and 6 months after surgery to assess early postoperative recovery and detect any new neurological deficits. At 12 months, a comprehensive clinical and radiological assessment was performed, including MRI and 6-vessel DSA. Long-term follow-up was conducted at regular intervals every three to five years and consisted of standardized clinical examination in conjunction with MRI and vascular imaging. Imaging studies were reviewed for the occurrence of new ischemic or hemorrhagic lesions, as well as for evaluation of bypass. Bypass function was primarily assessed using DSA. If DSA was not available, CT angiography or magnetic resonance angiography was used as an alternative. In cases of new or worsening neurological symptoms, additional imaging was obtained outside the scheduled follow-up intervals to differentiate ischemic from hemorrhagic events and to evaluate bypass integrity. Functional outcome at each follow-up time point was graded using the modified Rankin Scale (mRS), with the latest available assessment used for the primary outcome analysis.

### Statistical analysis

All analyses were performed in R version 4.3.2 (R Foundation for Statistical Computing, Vienna, Austria). The significance level was set at α=0.05. Continuous variables are presented as mean with standard deviation or median with interquartile range, and categorical variables as counts and percentages. Baseline characteristics were assessed descriptively without formal hypothesis testing, as group comparisons were not a prespecified objective. The primary outcome was functional status at the latest available follow-up, measured by the mRS and analyzed as a full ordered variable using proportional odds regression. The multivariable model included recorded sex, age, preoperative mRS, hypertension, diabetes mellitus, coronary artery disease, atrial fibrillation, chronic kidney disease, and peripheral artery disease, selected a priori on clinical grounds. Results are expressed as odds ratios (OR) with 95% confidence intervals (CI); ORs >1 indicate higher odds of a worse mRS category. Model assumptions, including proportional odds and collinearity, were assessed. For the subgroup analysis, CVRC was dichotomized as preserved (≥20%) or impaired (<20%). The CVRC subgroup analysis included only patients with available CVRC data without imputation. Robustness was assessed using a leave-one-covariate-out sensitivity analysis and by refitting after exclusion of influential observations identified using Cook’s distance. A detailed summary of the statistical analysis is provided in Supplement 1.

## RESULTS

Between February 2012 and March 2025, a total of 630 patients underwent EC-IC bypass at our institution. Of these, 275 patients were treated for ACVD and were considered eligible for further screening. After applying the predefined inclusion criteria, 135 patients were excluded due to incomplete follow-up, including domestic referrals (n=42), international referrals (n=43), and patients lost to follow-up (n=50). The remaining 140 patients constituted the final study cohort and were included in all full-cohort analyses (Figure 1).

**Figure 1.**
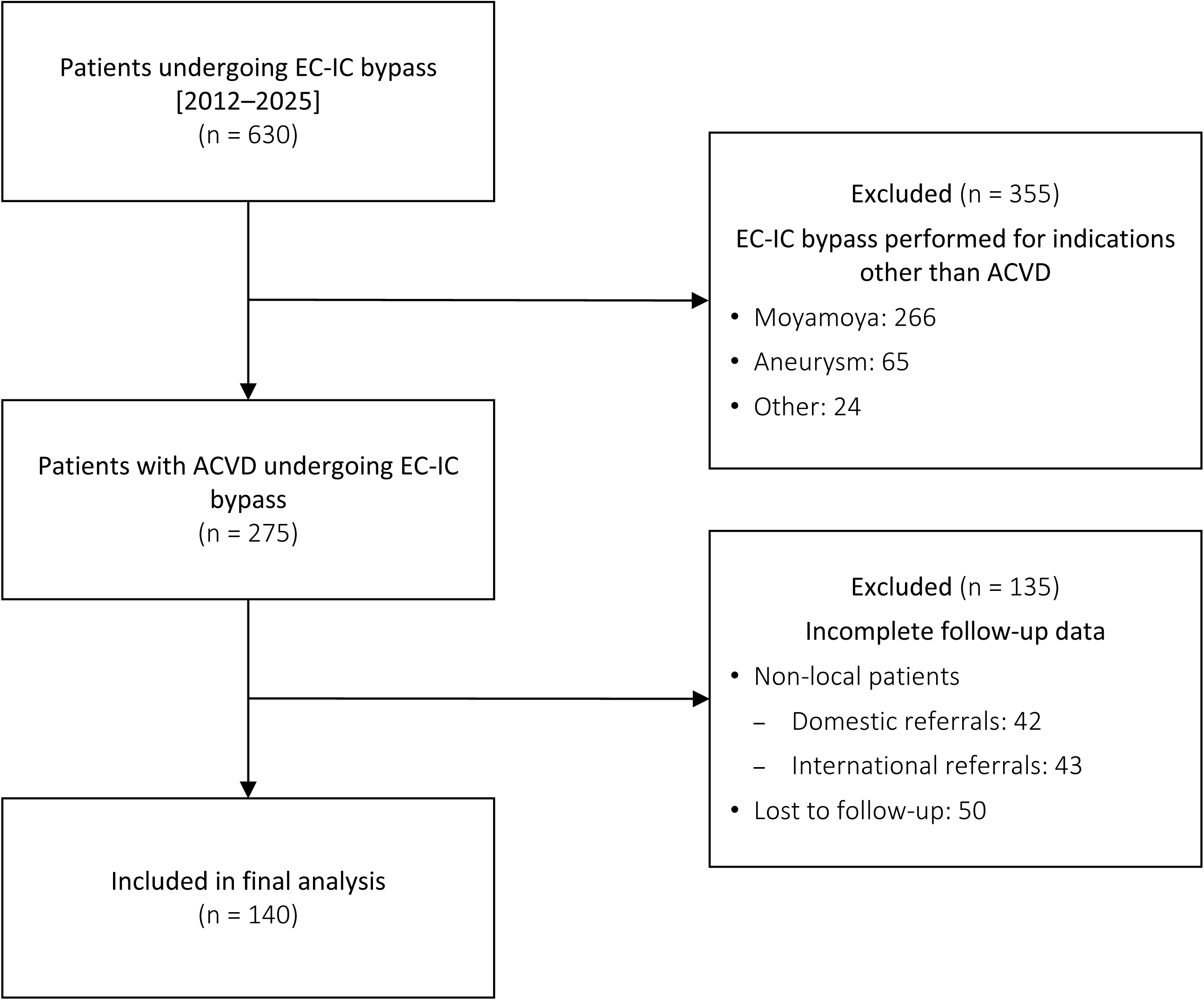
Patient selection flowchart. Flow diagram illustrating cohort selection from 630 patients undergoing EC-IC bypass (2012–2025) to the final study population of 140 patients with ACVD after exclusion of non-atherosclerotic indications and incomplete follow-up. ACVD = atherosclerotic cerebrovascular disease; EC-IC = extracranial–intracranial.

### Baseline characteristics

The cohort included 43 females (30.7%) and 97 males (69.3%), with baseline demographic and clinical characteristics summarized in Table 1. Age (57.8 ± 14.0 vs. 59.7 ± 9.1 years) and body mass index (BMI; 28.1 ± 5.6 vs. 28.1 ± 5.0 kg/m² in females and males, respectively) were comparable between sexes. Most patients in both groups were functionally independent, with 79.1% of females and 77.3% of males classified as mRS 0–2 preoperatively. A single ischemic event was the most frequent presentation in females (55.8%), whereas recurrent ischemic events were slightly more frequent in males (48.5%). Hypertension was the predominant comorbidity overall (81.4% vs. 87.6%), followed by hyperlipidemia (53.5% vs. 53.6%). Diabetes mellitus was more prevalent in males (38.1% vs. 27.9%), while other vascular comorbidities, including coronary artery disease (27.9% vs. 24.7%), chronic kidney disease (9.3% vs. 5.2%), and atrial fibrillation (4.7% vs. 4.1%), were similarly distributed. The majority of patients received single antiplatelet therapy and statins in both groups.

**Table 1:** Baseline demographic and clinical characteristics of patients undergoing superficial temporal artery – middle cerebral artery (STA-MCA) bypass surgery, stratified by sex. Continuous variables are presented as mean ± standard deviation and median (interquartile range), and categorical variables as absolute (relative) frequencies. COPD = chronic obstructive pulmonary disease; SD = standard deviation.

|  | All<br>(n=140) | Female<br>(n=43) | Male<br>(n=97) |
| --- | --- | --- | --- |
| **Demographics** |  |  |  |
| Age, years |  |  |  |
| Mean ± SD | 59.2 ± 10.9 | 57.8 ± 14.0 | 59.7 ± 9.1 |
| Median (IQR) | 60 (53-67) | 59 (46-68) | 60 (55-66) |
| Body mass index, kg/m<sup>2</sup> |  |  |  |
| Mean ± SD | 28.1 ± 5.1 | 28.1 ± 5.6 | 28.1 ± 5.0 |
| Median (IQR) | 28.0 (24.2-31.2) | 27.3 (24.1-30.6) | 28.0 (25.0-31.2) |
| **Onset symptoms, n (%)** |  |  |  |
| Single ischemic event | 69 (49.3%) | 24 (55.8%) | 45 (46.4%) |
| Recurrent ischemic events | 66 (47.1%) | 19 (44.2%) | 47 (48.5%) |
| Other / asymptomatic | 5 (3.6%) | 0 (0%) | 5 (5.2%) |
| **Comorbidities, n (%)** |  |  |  |
| Hypertension | 120 (85.7%) | 35 (81.4%) | 85 (87.6%) |
| Diabetes mellitus | 49 (35.0%) | 12 (27.9%) | 37 (38.1%) |
| Hyperlipidemia | 75 (53.6%) | 23 (53.5%) | 52 (53.6%) |
| Smoking | 37 (26.4%) | 9 (20.9%) | 28 (28.9%) |
| COPD | 8 (5.7%) | 3 (7.0%) | 5 (5.2%) |
| Coronary artery disease | 36 (25.7%) | 12 (27.9%) | 24 (24.7%) |
| Peripheral artery disease | 21 (15.0%) | 9 (20.9%) | 12 (12.4%) |
| Atrial fibrillation | 6 (4.3%) | 2 (4.7%) | 4 (4.1%) |
| Chronic kidney disease | 9 (6.4%) | 4 (9.3%) | 5 (5.2%) |
| **Medications, n (%)** |  |  |  |
| Antiplatelet |  |  |  |
| Single | 105 (75.0%) | 31 (72.1%) | 74 (76.3%) |
| Double | 32 (22.9%) | 12 (27.9%) | 20 (20.6%) |
| None | 3 (2.1%) | 0 (0%) | 3 (3.1%) |
| Statin | 113 (80.7%) | 32 (74.4%) | 81 (83.5%) |
| Anticoagulation | 5 (3.6%) | 1 (2.3%) | 4 (4.1%) |
| **Modified Rankin Scale, n (%)** |  |  |  |
| 0 | 30 (21.4%) | 9 (20.9%) | 21 (21.6%) |
| 1 | 52 (37.1%) | 16 (37.2%) | 36 (37.1%) |
| 2 | 27 (19.3%) | 9 (20.9%) | 18 (18.6%) |
| 3 | 19 (13.6%) | 3 (7.0%) | 16 (16.5%) |
| 4 | 11 (7.9%) | 5 (11.6%) | 6 (6.2%) |
| 5 | 1 (0.7%) | 1 (2.3%) | 0 (0%) |

### Disease characteristics and vascular pathology

Sex-specific patterns in cerebrovascular disease distribution and hemodynamic characteristics are summarized in Table 2. Females more frequently presented with multivessel involvement (65.1% vs. 47.4%), particularly combined internal carotid artery (ICA) and anterior circulation disease (27.9% vs. 11.3%). In contrast, isolated ICA disease was more common in males (43.3% vs. 16.3%). Occlusive disease predominated overall but was more frequent in males (79.4% vs. 60.5%), whereas stenotic lesions were relatively more common in females (39.5% vs. 20.6%). Bilateral hemispheric involvement was higher in males (42.3% vs. 14.0%), while females more often demonstrated left-sided predominance (48.8% vs. 34.0%). CVRC was available in 123 patients (87.9%). Impaired CVRC (>0 to <20%) was more frequent in males (62.9% vs. 41.9%), whereas mildly reduced CVRC (20–40%) was more frequent in females (18.6% vs. 2.1%). Exhausted CVRC (0%) was similarly distributed (23.3% vs. 22.7%).

**Table 2:** Disease characteristics and operative details of patients undergoing superficial temporal artery – middle cerebral artery (STA-MCA) bypass surgery, stratified by sex. Continuous variables are presented as mean ± standard deviation and median (interquartile range), and categorical variables as absolute (relative) frequencies. ICA = internal carotid artery; CCA = common carotid artery; CVRC = cerebrovascular reserve capacity; IQR = interquartile range; MCA = middle cerebral artery; mRS = modified Rankin Scale; SD = standard deviation.

|  | All<br>(n=140) | Female<br>(n=43) | Male<br>(n=97) |
| --- | --- | --- | --- |
| **Affected vessel pattern, n (%)** |  |  |  |
| CCA isolated | 5 (3.6%) | 1 (2.3%) | 4 (4.1%) |
| ICA isolated | 49 (35.0%) | 7 (16.3%) | 42 (43.3%) |
| MCA isolated | 12 (8.6%) | 7 (16.3%) | 5 (5.2%) |
| Multivessel Disease | 74 (52.9%) | 28 (65.1%) | 46 (47.4%) |
| Bilateral ICA | 12 (8.6%) | 2 (4.7%) | 10 (10.3%) |
| ICA + anterior circulation | 23 (16.4%) | 12 (27.9%) | 11 (11.3%) |
| ICA + posterior circulation | 15 (10.7%) | 5 (11.6%) | 10 (10.3%) |
| Other multivessel patterns | 24 (17.1%) | 9 (20.9%) | 15 (15.5%) |
| **Vessel status, n (%)** |  |  |  |
| Stenosis | 37 (26.4%) | 17 (39.5%) | 20 (20.6%) |
| Occlusion | 103 (73.6%) | 26 (60.5%) | 77 (79.4%) |
| **Affected hemisphere, n (%)** |  |  |  |
| Left | 54 (38.6%) | 21 (48.8%) | 33 (34.0%) |
| Right | 39 (27.9%) | 16 (37.2%) | 23 (23.7%) |
| Bilateral | 47 (33.6%) | 6 (14.0%) | 41 (42.3%) |
| **Perfusion status (CVRC), n (%)** |  |  |  |
| Normal, (>40% CVRC) | 2 (1.4%) | 0 (0.0%) | 2 (2.1%) |
| Slightly reduced, (20-40% CVRC) | 10 (7.1%) | 8 (18.6%) | 2 (2.1%) |
| Reduced, (>0 to <20% CVRC) | 79 (56.4%) | 18 (41.9%) | 61 (62.9%) |
| Exhausted, (0% CVRC) | 32 (22.9%) | 10 (23.3%) | 22 (22.7%) |
| Not available | 17 (12.1%) | 7 (16.3%) | 10 (10.3%) |
| **Hospital stay, days** |  |  |  |
| Mean ± SD | 10.6 ± 7.2 | 11.5 ± 8.4 | 10.3 ± 6.6 |
| Median (IQR) | 9 (7-14) | 10 (6.7-14) | 8 (7-14) |
| **mRS at discharge, n (%)** |  |  |  |
| 0 | 21 (15.0%) | 5 (11.6%) | 16 (16.5%) |
| 1 | 70 (50.0%) | 19 (44.2%) | 51 (52.6%) |
| 2 | 22 (15.7%) | 9 (20.9%) | 13 (13.4%) |
| 3 | 12 (8.6%) | 3 (7.0%) | 9 (9.3%) |
| 4 | 12 (8.6%) | 5 (11.6%) | 7 (7.2%) |
| 5 | 0 (0.0%) | 0 (0.0%) | 0 (0.0%) |
| 6 | 3 (2.1%) | 2 (4.7%) | 1 (1.0%) |

### Perioperative course and follow-up outcomes

The 30-day stroke rate was 9.3% in females and 1.0% in males (Table 3). Additional perioperative events within 30 days included hemorrhagic events and other complications, occurring exclusively in males (each 2/97, 2.1%). Hospital stay was slightly longer in females (median 10 vs. 8 days). Mean follow-up duration was comparable between females (24.1 ± 24.5 months) and males (23.1 ± 24.4 months). New neurological symptoms during follow-up were recorded at similar rates in both sexes (18.6% vs. 19.6%). Ischemic events predominated in females (16.3% vs. 7.2%), while hemorrhagic events occurred exclusively in males (5.2% vs. 0%), of which four were parenchymal intracerebral hemorrhages and one was a subdural hematoma. Bypass occlusion was rare and observed only in males (n=2). At latest follow-up, functional outcome differed between sexes, with a higher proportion of males having an mRS of 0–1 (62.9% vs. 41.8%). Conversely, an mRS ≥2 was more frequent in females (58.1% vs. 37.1%). Mortality (mRS 6) was observed in 7.0% of females and 3.1% of males. Among the six patients with mRS 6 (three males, three females), all male deaths were due to intracerebral bleeding (one perioperative, two post-discharge), whereas all female deaths resulted from ischemic stroke (two perioperative, one post-discharge), consistent with the predominance of ischemic events in females and hemorrhagic events in males observed throughout the follow-up period. Notably, most patients with severe postoperative disability (mRS ≥4) already had substantial functional impairment before surgery. Only three patients with a preoperative mRS of 1–2 progressed to an mRS ≥4, all of whom experienced postoperative infarction.

**Table 3:** Follow-up characteristics and clinical outcomes after superficial temporal artery – middle cerebral artery (STA-MCA) bypass surgery, stratified by sex. Continuous variables are presented as mean ± standard deviation and median (interquartile range), and categorical variables as absolute (relative) frequencies. FU = follow-up; IQR = interquartile range; SD = standard deviation.

|  | All<br>(n=140) | Female<br>(n=43) | Male<br>(n=97) |
| --- | --- | --- | --- |
| **Mean time until latest available FU** |  |  |  |
| Months, mean ± SD | 23.4 ± 24.3 | 24.1 ± 24.5 | 23.1 ± 24.4 |
| Months, median (IQR) | 13.5 (5-37) | 14 (3.5-40) | 13 (6.0-32) |
| **30-day stroke rate, n (%)** | 5 (3.6%) | 4 (9.3%) | 1 (1.0%) |
| **30-day perioperative events, n (%)** |  |  |  |
| Hemorrhagic event | 2 (1.4%) | 0 (0%) | 2 (2.1%) |
| Other | 2 (1.4%) | 0 (0%) | 2 (2.1%) |
| **New symptoms at FU, n (%)** | 27 (19.3%) | 8 (18.6%) | 19 (19.6%) |
| Ischemic symptoms | 14 (10.0%) | 7 (16.3%) | 7 (7.2%) |
| Hemorrhagic symptoms | 5 (3.6%) | 0 (0%) | 5 (5.2%) |
| Others | 8 (5.7%) | 1 (2.3%) | 7 (7.2%) |
| **Bypass occlusion, n (%)** | 2 (1.4%) | 0 (0%) | 2 (2.1%) |
| **Modified Rankin Scale, n (%)** |  |  |  |
| 0 | 43 (30.7%) | 9 (20.9%) | 34 (35.1%) |
| 1 | 36 (25.7%) | 9 (20.9%) | 27 (27.8%) |
| 2 | 28 (20.0%) | 12 (27.9%) | 16 (16.5%) |
| 3 | 17 (12.1%) | 4 (9.3%) | 13 (13.4%) |
| 4 | 7 (5.0%) | 4 (9.3%) | 3 (3.1%) |
| 5 | 3 (2.1%) | 2 (4.7%) | 1 (1.0%) |
| 6 | 6 (4.3%) | 3 (7.0%) | 3 (3.1%) |

### Regression analysis and sensitivity analysis

In the univariable analysis, preoperative functional status was the strongest predictor of outcome, with each increase in baseline mRS associated with a more than four-fold higher odds of worse follow-up mRS (OR 4.14, 95% CI 2.98–5.89, p<0.001; Figure 2). Female sex was associated with significantly worse functional outcome (OR 2.16, 95% CI 1.13–4.15, p=0.020), alongside age (OR 1.03 per year, 95% CI 1.00–1.06, p=0.043), coronary artery disease (OR 2.20, 95% CI 1.12–4.34, p=0.023), and anticoagulation therapy (OR 5.67, 95% CI 1.28–24.74, p=0.019). After multivariable adjustment for clinically relevant covariates, female sex remained independently associated with worse functional outcome (OR 2.59, 95% CI 1.28–5.30, p=0.008; Figure 3). Preoperative mRS remained the strongest predictor of outcome (OR 4.30, 95% CI 3.07–6.18, p<0.001), while peripheral artery disease, chronic kidney disease, and hypertension showed consistent but non-significant trends toward worse outcome. The 30-day stroke rate and perioperative events could not be included in the regression analysis given the low absolute event count. Sensitivity analyses confirmed the stability of the association between female sex and worse functional outcome across all leave-one-covariate-out iterations (Supplement 2). The effect size remained consistent after sequential exclusion of individual covariates and remained statistically robust (OR range approximately 2.4–2.7). In an exploratory analysis, exclusion of observations identified by Cook’s distance did not materially alter the results, with persistent significance and similar effect magnitude.

**Figure 2.**
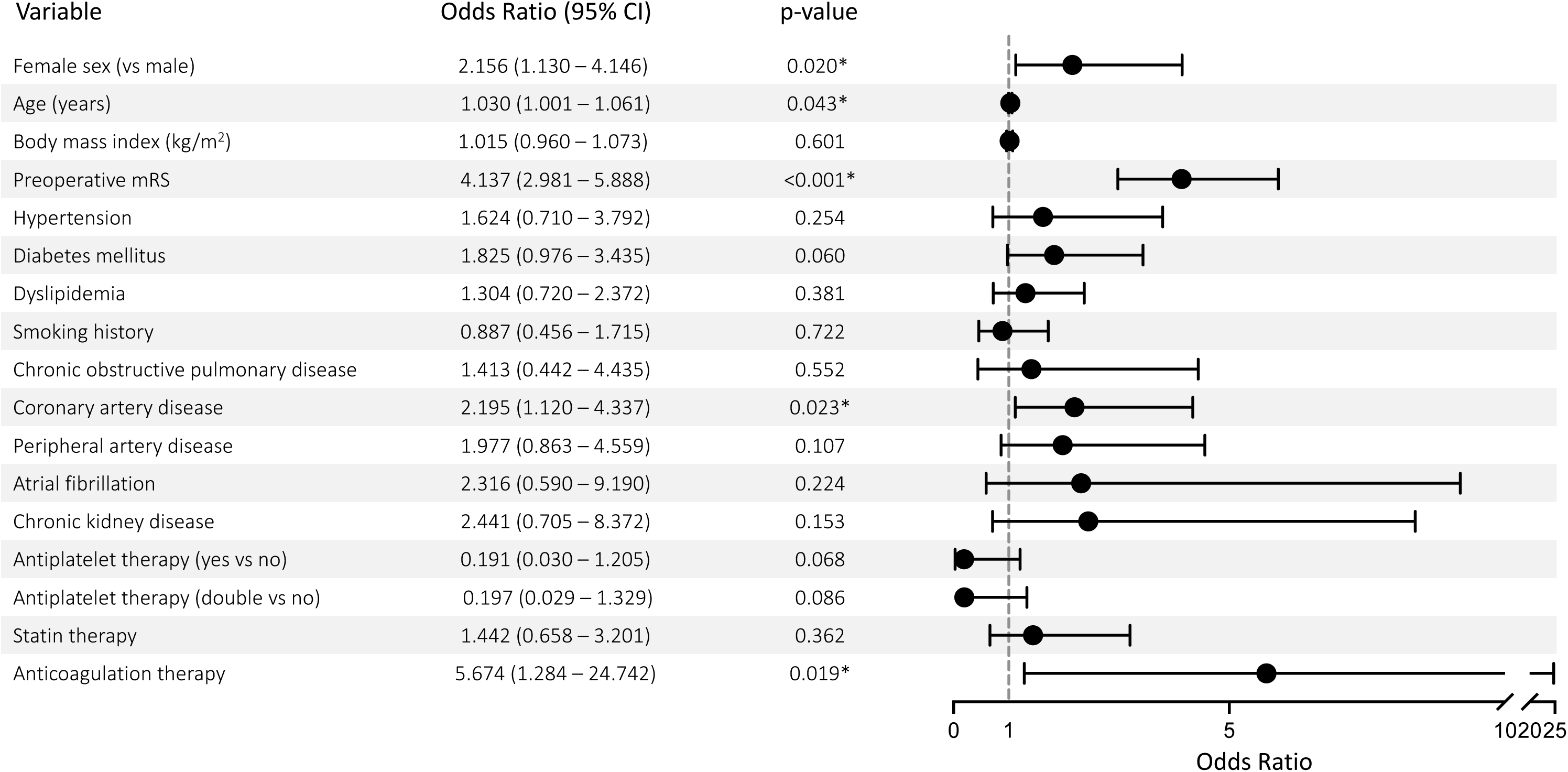
Univariable ordinal proportional odds regression analysis of variables associated with functional outcome. Forest plot showing odds ratios (OR) with 95% confidence intervals (CI) from univariable ordinal proportional odds regression for prespecified variables in relation to modified Rankin Scale (mRS) at latest follow-up. CI = confidence interval; mRS = modified Rankin Scale; OR = odds ratio. OR >1 indicates higher odds of a worse mRS category; *p<0.05.

**Figure 3.**
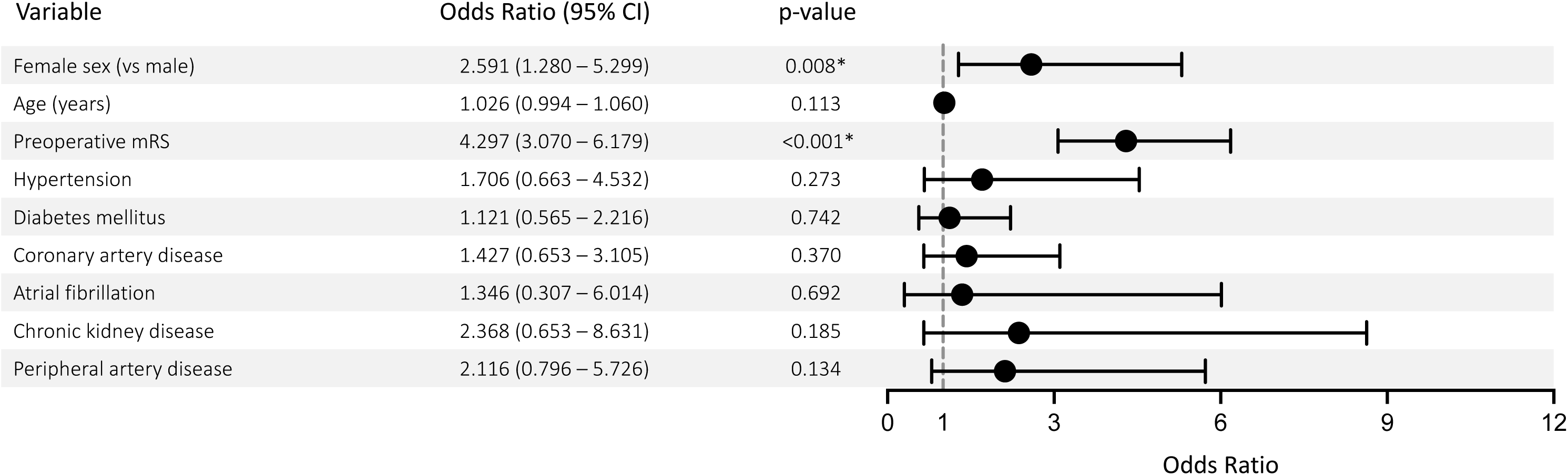
Multivariable ordinal proportional odds regression analysis of variables associated with functional outcome. Forest plot displaying adjusted odds ratios (OR) with 95% confidence intervals (CI) from multivariable ordinal proportional odds regression including prespecified covariates for modified Rankin Scale (mRS) at latest follow-up. CI = confidence interval; mRS = modified Rankin Scale; OR = odds ratio. OR >1 indicates higher odds of a worse mRS category; *p<0.05.

### CVRC subgroup analysis

Among patients with available cerebrovascular reserve data (n=123), the majority demonstrated markedly impaired or exhausted cerebrovascular reserve capacity (90.2%). For analysis, CVRC was dichotomized into preserved (≥20%) and impaired (<20%) reserve. Within this subgroup, female sex remained independently associated with worse functional outcome (OR 2.42, 95% CI 1.09–5.45, p=0.031), with preoperative mRS again the strongest predictor (OR 4.30, 95% CI 2.98–6.41, p<0.001). No statistically significant association between cerebrovascular reserve status and outcome was detected in adjusted models (preserved vs. impaired CVRC: OR 0.80, 95% CI 0.24–2.70, p=0.721; Table 4).

**Table 4:** Multivariable ordinal proportional odds regression analysis in the cerebrovascular reserve capacity subgroup. Results are presented as odds ratios with 95% confidence intervals (CI) and corresponding p-values. An OR >1 indicates higher odds of being in a worse modified Rankin Scale category. *p<0.05. CVRC = cerebrovascular reserve capacity.

|  | Odds Ratio | 95% Confidence Interval | p-value |
| --- | --- | --- | --- |
| Female sex | 2.423 | 1.090–5.449 | 0.0305* |
| Age | 1.016 | 0.981–1.054 | 0.3778 |
| Preoperative mRS | 4.303 | 2.977–6.411 | <0.0001* |
| Hypertension | 1.672 | 0.623–4.692 | 0.3156 |
| Diabetes mellitus | 1.157 | 0.547–2.430 | 0.7012 |
| Coronary artery disease | 1.421 | 0.616–3.250 | 0.4059 |
| Atrial fibrillation | 1.479 | 0.337–6.635 | 0.6022 |
| Chronic kidney disease | 2.419 | 0.563–10.067 | 0.2223 |
| Peripheral artery disease | 2.18 | 0.776–6.200 | 0.1388 |
| Preserved CVRC | 0.802 | 0.235–2.695 | 0.7211 |

## DISCUSSION

In this single-center retrospective cohort study, we examined the association of biological sex with functional outcome following STA-MCA bypass in 140 patients with ACVD. Female sex was independently associated with worse functional outcome across all analytical approaches. An important observation is that women also demonstrated a distinctly different vascular phenotype. Women more frequently presented with multivessel disease and stenotic lesions, whereas isolated ICA occlusion predominated in men. Although multivariable adjustment supported an independent association between sex and outcome, the more complex vascular disease pattern observed in women may nevertheless biologicaly contribute to the observed outcome differences and should be considered when interpreting the present findings.

### Baseline characteristics and disease morphology

The demographic and comorbidity profile of the present cohort reflects a real-world bypass population that differs in relevant aspects from the pivotal randomized trials. The mean age of 59.2 years was broadly similar to that in COSS and older than that in CMOSS^13,14^. The female proportion of 30.7% is comparable to that in COSS (approximately 30% female) and CMOSS (approximately 20% female)^13,14^, neither of which performed sex-stratified outcome analyses, a gap the present study directly addresses. Hypertension was the dominant comorbidity in both sexes (81.4% female, 87.6% male), consistent with the risk factor profiles of both trials^13,14^. Diabetes mellitus was more prevalent in males (38.1% vs. 27.9%), a distribution directionally consistent with the male-predominant cohorts of the landmark trials^13,14^. Crucially, age, BMI, hyperlipidemia, and antiplatelet or statin use were comparable between sexes, suggesting that the observed outcome disparity is unlikely to be attributable to an imbalanced cardiovascular risk profile alone. Females more commonly presented with multivessel (65.1% vs. 47.4%) and stenotic disease (39.5% vs. 20.6%), while males showed a higher proportion of isolated ICA occlusion (43.3% vs. 16.3%), a distribution consistent with greater overall disease complexity in females. Nevertheless, the overall disease burden in our cohort exceeded that of the landmark trials, consistent with our previously reported trend in this population^18^.

### Sex-related outcome disparities in cerebrovascular disease

Despite growing interest in sex-specific outcomes in cerebrovascular disease, corresponding data in bypass surgery remain scarce. The only prior sex-stratified analysis in this surgical context, by McGuire et al., was limited to a mixed-diagnosis cohort and focused on bypass patency rather than postoperative functional recovery, leaving this dimension unaddressed^12^. The present study therefore represents, to our knowledge, the first to assess functional outcome as the primary endpoint in a sex-stratified analysis of direct STA-MCA bypass for ACVD. In the broader cerebrovascular literature, biological sex has repeatedly been associated with procedural risk. In carotid revascularization, higher periprocedural complication rates in women have been reported in several cohorts^6,7^, though not consistently^19,20^. The prespecified sex-stratified analysis of CREST demonstrated a significantly higher periprocedural event rate in women assigned to stenting versus endarterectomy (HR 1.84, 95% CI 1.01–3.37), while pooled analyses across EVA-3S, SPACE, ICSS, and CREST reported no consistent sex differences in outcomes, limited by inter-trial heterogeneity^3,21^. From the perspective of acute stroke management, female sex has been associated with less favorable functional outcomes following intravenous thrombolysis for acute ischemic stroke^1,4^ and sex-specific differences in antiplatelet treatment response have been reported in secondary prevention, with dual antiplatelet therapy reducing stroke recurrence in males but not females^22^. The pathophysiological basis of these disparities remains incompletely understood. Beyond disease burden and perioperative events, sex-related differences in systemic atherosclerotic disease expression, including plaque composition and endothelial function, may further modulate procedural risk and recovery in ways not captured by standard preoperative assessment, although their specific role in bypass surgery remains to be established.^7,23,24^ Whether the observed association reflects biological differences per se or differences in disease phenotype and vascular complexity cannot be fully determined within the present retrospective design.

### Role of preoperative functional status

A central interpretive issue in sex–outcome research is separating the association with biological sex from pre-existing functional vulnerability. The OXVASC study demonstrated that worse outcomes in women with vascular diseases were largely explained by higher premorbid mRS, with the sex difference disappearing after adjustment^25^. In post-thrombectomy analyses, pre-stroke functional status consistently emerges as one of the strongest determinants of 90-day independence, alongside age and stroke severity, underscoring the dominant role of baseline neurological reserve in outcome prediction^5^. In contrast to most prior observational series^26–28^, we explicitly adjusted for preoperative mRS as a continuous predictor, and the association not only persisted but increased in magnitude compared with univariable analysis (OR 2.16 vs. 2.59). The increase in the estimate after adjustment may reflect negative confounding or statistical suppression and should not be interpreted as evidence for a biological mechanism. Notably, within this multivariable framework, preoperative mRS nonetheless emerged as the strongest independent predictor of outcome, exceeding the effect of sex and other covariates. This finding has potential clinical implications.

Rather than advocating different surgical thresholds, however, it underscores the importance of incorporating biological sex into stroke prevention strategies and supports considering sex as an additional variable in future risk stratification models and prospective studies aimed at individualized treatment.

### CVRC: selection tool versus outcome determinant

CVRC assessed by acetazolamide-stimulated SPECT or PET provided the hemodynamic rationale for patient selection in this cohort, but no statistically significant independent association with functional outcome was detected. The lack of a statistically significant association between preoperative CVRC and postoperative functional outcome should be interpreted cautiously. Nearly all patients included fulfilled contemporary hemodynamic indications for bypass surgery, resulting in limited variability of CVRC within the study cohort. Therefore, these findings should not be interpreted as questioning the value of CVRC for patient selection, but rather indicate that within a highly selected population, preoperative CVRC alone was not shown to explain postoperative recovery. Future studies evaluating postoperative improvement of cerebrovascular reserve rather than baseline impairment alone may provide further mechanistic insight.

Interestingly, females presented with less severe CVRC impairment despite more complex disease distribution, a paradox that may reflect sex-specific differences in collateral physiology and microvascular resilience not fully captured by macro-hemodynamic reserve metrics. Clinical thrombectomy studies have similarly reported superior collateral flow in women despite less favorable functional outcomes^29^. Experimental studies suggest sex-related differences in ischemic tolerance that are not fully explained by native leptomeningeal collateral extent or remodeling.^30^ Separate preclinical work has demonstrated that collateralization in chronic cerebral ischemia can be therapeutically enhanced,^31,32^ highlighting the adaptive potential of the cerebral collateral circulation. Together, these findings provide a rationale for further investigation of sex-specific neurovascular adaptation. These observations raise the possibility that sex-specific differences in collateral physiology and ischemic adaptation may partially compensate for more extensive disease in females, but do not appear to translate into superior functional recovery after bypass. Postoperative functional recovery may therefore depend not only on baseline macro-hemodynamic reserve, but also on the capacity for sex-specific neurovascular adaptation following revascularization.

### Perioperative and follow-up event pattern

The overall 30-day symptomatic ischemic stroke rate was 3.6%. Direct comparison with the surgical arms of COSS and CMOSS is limited because their perioperative endpoints differed from the endpoint used here^13,14^. However, a marked sex-specific variation was observed, with a 30-day stroke rate of 9.3% in females compared with 1.0% in males. Interestingly, female patients experienced predominantly ischemic complications whereas hemorrhagic complications occurred exclusively in men. This directional imbalance may partly reflect the greater burden of multivessel and stenotic disease in females, although the small absolute event count precludes definitive conclusions. Beyond the perioperative period, ischemic events continued to account for the majority of new neurological symptoms in females (16.3% vs. 7.2%), while males demonstrated a more heterogeneous distribution including hemorrhagic complications (5.2% vs. 0%). Taken together, these findings point to a distinct clinical phenotype in female bypass patients, characterized by greater perioperative stroke susceptibility and an ischemia-predominant event profile, the mechanistic basis of which warrants dedicated investigation.

### Limitations

This study is retrospective and from a single center, with an inherent risk of residual confounding and selection bias. Almost half of potentially eligible patients with ACVD were excluded because of incomplete follow-up, which may have introduced additional selection bias. The primary outcome was assessed at the latest available follow-up, which varied between patients. The generalizability of these findings is limited by the study design, and external validity in other patient populations and healthcare settings remains to be established. The smaller female sample size (n=43) limits the precision of sex-stratified estimates. CVRC data were unavailable in some patients, limiting precision in the subgroup analysis. Psychosocial factors such as postoperative depression and social support, along with unmeasured variables including menopausal status, hormonal therapy, frailty indices, and cognitive reserve, were not systematically captured but may influence functional recovery differently in men and women. Because disease morphology differed between sexes and was not included in the multivariable model, residual confounding by anatomical complexity cannot be excluded. Despite these limitations, the consistency of the association across sensitivity analyses and in the CVRC subgroup supports the robustness of the primary finding.

## CONCLUSION

Female sex was independently associated with worse functional outcome after STA-MCA bypass for ACVD in this observational cohort. Whether this reflects intrinsic biological differences, more complex cerebrovascular disease, or both, requires confirmation in prospective multicenter studies.

## DECLARATIONS

### Ethical approval

All procedures performed in this study involving human participants were in accordance with the ethical standards of the institutional and/or national research committee (ethics committee of the Charité *–* Universitätsmedizin Berlin, Germany; EA2/139/12 and EA2/178/18) and with the Declaration of Helsinki (1964) and its later amendments or comparable ethical standards.

### Informed consent

Formal consent for retrospective analysis was waived under ethical approvals EA2/139/12 and EA2/178/18.

### Consent for publication

Included within the Ethics approval of the Charité *–* Universitätsmedizin Berlin, Germany (EA2/139/12 and EA2/178/18), granted to the authors of the study.

### Data availability

Available upon reasonable request from the corresponding author (NH). Access is restricted to protect patient confidentiality. Extended descriptive statistics are available through the statistical analysis report (Supplement 1). Analysis code is available at https://github.com/iqbal-ma/Impact-of-sex-on-bypass-outcome.

### Author contributions

Conception and design: NH. Data acquisition: MI, JA, LW, PV, GA, NH. Data interpretation: all authors. Statistical analysis: MI, EDS, NH. Drafting of the manuscript: MI. Critical revision of the manuscript for important intellectual content: all authors.

### Use of large language models

Claude (Anthropic PBC, San Francisco, CA, USA) was used to enhance readability based on grammar, spelling and syntax. All scientific content, data interpretation, and intellectual conclusions are solely the work of the authors.

## Acknowledgments

None.

## ABBREVIATIONS

ACVD: Atherosclerotic cerebrovascular disease
BMI: Body mass index,
CI: Confidence interval,
CMOSS: Chinese Multicenter Occlusion Surgery Study
COSS: Carotid Occlusion Surgery Study
CT: Computed tomography
CVRC: Cerebrovascular reserve capacity
DSA: Digital subtraction angiography
EC-IC: Extracranial–intracranial,
ICA: Internal carotid artery,
MRI: Magnetic resonance imaging
mRS: Modified Rankin Scale
OR: Odds ratio
PET: Positron emission tomography
SPECT: Single-photon emission computed tomography
STA-MCA: Superficial temporal artery to middle cerebral artery
TIA: Transient ischemic attack.

## SUPPLEMENT LEGENDS

**Supplement 1. Detailed statistical analysis report.** Comprehensive statistical analysis report describing data preprocessing, model specification, assessment of proportional odds and collinearity assumptions, sensitivity analyses, handling of influential observations, and additional descriptive analyses supporting the multivariable ordinal regression models for functional outcome.

**Supplement 2. Leave-one-covariate-out sensitivity analysis of the theory-driven multivariable ordinal proportional odds regression.** Plot showing the stability of the female-sex estimate in the theory-driven multivariable model for modified Rankin Scale (mRS) at latest follow-up after sequential exclusion of individual covariates. Effect estimates (odds ratios with 95% confidence intervals) are displayed for each iteration. CI = confidence interval; mRS = modified Rankin Scale; OR = odds ratio. OR >1 indicates higher odds of a worse mRS category; *p<0.05.

